# Incubation period and serial interval of Covid-19 in a chain of infections in Bahia Blanca (Argentina)

**DOI:** 10.1101/2020.06.18.20134825

**Authors:** Valentina Viego, Milva Geri, Juan Castiglia, Ezequiel Jouglard

## Abstract

**Objective:** To estimate the incubation period and the serial interval of Covid-19 from a sample of symptomatic patients in Bahia Blanca city.

**Methods:** We collected dates of illness onset for primary cases (infectors) and secondary cases (infectees) for the first 18 secondary patients infected with SARS-Cov-2 in Bahia Blanca (Argentina). We ranked the fiability of the data depending upon certainty about the identification of the infector and the date of exposition to infector.

The sample has some missing values. In the case of incubation, as 3 patients were infected by other household members, we only have 15 observations with an observed date of exposition. For the estimation of serial interval, one patient became ill from close contact with an asymptomatic infectious. Also, estimations of both the incubation period and the serial interval were carried using the full sample and a subsample with higher certainty about the transmissor and date of exposition. By the time the dataset was prepared all infectors were recovered so estimations do need to take into account right censoring.

**Results:** The mean incubation period for symptomatic patients is 7.9 days (95% CI: 4.6, 11.1) considering the sample of 15 cases patients and 7.5 days (95% CI: 4.1, 10.9) if the sample is restricted to the most certain cases (n=12). The median is 6.1 (95% CI: 4.1, 9.2) and 5.8 (95% CI: 3.6, 9.3) respectively. Moreover, 97.5% of symptomatic cases will develop symptoms afert 13.6 days from exposition (95% CI 10.7, 16.5).

The point estimation for the mean serial interval is 6.8 days (95% CI: 4.0-9.6). Considering only the most certain pairs, the mean serial interval is estimated at 5.5 days (95% CI: 2.8, 8.1). The estimated median serial intervals were 5.2 (95% CI: 3.0, 8.1) and 4.1 (95% CI: 2.0, 6.9) days respectively.

**Conclusions:** Evidence from Bahia Blanca (Argentina) suggests that the median and mean serial interval of Covid-19 is shorter than the incubation period. This suggests that a pre-symptomatic transmission is not negligible. Comparisons with foreign estimates show that incubation period and serial interval could be longer in Bahia Blanca city than in other regions. That poses a signal of opportunity to attain more timely contact tracing and effective isolation.

**Highlights:** We estimate the incubation period in a sample of 15 symptomatic patients with Covid-19 in Bahia Blanca city (Argentina).

We estimate the serial interval for Covid-19 infections in a sample of 17 infector-infectee pairs detected in Bahia Blanca city (Argentina).

The median serial interval is lower to the median incubation period, suggesting a transmission is taking place also during the pre-symptomatic phase.

The incubation period and serial interval of Covid-19 in Bahía Blanca city seem to take more days than in Asian regions. This finding slows down the pace of health assistance to patients (conditional to public interventions).

Longer serial intervals help in tracing contacts and show relative slow turnover of case generations. At the same time, if symptoms take longer time to emerge, long serial intervals may also increase the reproductive number if contact tracing and effective isolation measures are placed untimely.

## Introduction

In Argentina the first case of the epidemic of Covid-19 was reported on March 3, 2020. As the WHO declared the disease as pandemic by March 11, 2020, health authorities decided to pose a mandatory lockdown since March 20, 2020 in order to prevent rapid propagation of infections and prepare the expansion of health infrastructure. After April 6, 2020, the government adopted a sequential easing of movements for selected economic activities combined with social distance measures, such as wearing masks and preserve the required distance between people.

Even though it is known that isolation and interventions that limit populations movements and contacts curb the spread, health authorities still do not know how the virus is spreading within the national borders and in specific territories, like towns, cities or densely populated areas. Also, as testing is limited, the proportion of asymptomatic is not known nor if contagion takes place during the pre-symptomatic phase of the disease. Besides the basic figures about confirmed cases, recovered or deaths, the development of local epidemiological indicators is still weak.

In infectious diseases, one of the key indicators is the serial interval (SI from now on), defined as the time from illness onset in infector (index or primary case) to illness onset in infectee (secondary case). This indicator contributes to the understanding of the transmissibility of the disease (Fine 2003). Actually the SI is widely used to compute the effective reproductive number, that is the average number of secondary infections caused by each infector (Wallinga & Lipsitch 2007). Estimates of the SI can only be obtained by linking the dates of illness onset between infector-infectee pairs. Thus, the main source of information emerges from clusters of infections.

Up to date, available figures of SI for Covid-19 come from a few regions, the majority belonging to regions with an early outbreak (see Table 1). As Peak et al (2020) point out, more data about the serial interval is needed to evaluate interventions (contact tracing, selective quarantines, etc). If SI is overestimated quarantine interventions may be excessive. On the contrary, if SI is underestimated, interventions may be insufficient to curb the spread. At the same time, if SI is relatively short, symptomatic cases will emerge rapidly and health authorities will need to focus efforts in testing infrastructure and coordination. If SI takes longer it may be a signal of low asymptomatic transmission and less pressure on testing inputs.

**Table 1.** Serial interval estimations for Covid-19. International evidence.

| Study | Region | Number of pairs | Median | Mean |
| --- | --- | --- | --- | --- |
| Li et al (2020) | Wuhan | 6 | not reported | 7.5<br>[5.3-19.0] |
| Ki (2020) | S. Korea | 12 | 4.0 | 6.6<br>[3.0-9.0] |
| Du et al (2020) | China excl Hubei | 468 | not reported | 3.96<br>[3.53-4.39] |
| Zhang et al (2020) | China excl Hubei | 35 | not reported | 5.1<br>[1.3-11.6] |
| Zhou et al (2020) | China excl Hubei | 71 | 4.0 | 4.27<br>(3.44) |
| Zhao et al (2020) <sup>U</sup> | Hong Kong | 21 | not reported | 4.4<br>[2.9-6.7] |
| Nishiura et al (2020) | Mainly Asian regions and cases reported in Germany | 28* | 4.0 | 4.7<br>[3.7-6.0] |
| Cereda et al (2020) <sup>U</sup> | Lombardy (Italy) | 90 | not reported | 6.6<br>[0.7-19] |
| Mettler & Maathuis (2020) <sup>U</sup> | S. Korea | 19 | not reported | 3.58<br>[2.62-4.53] |

standard deviations in parenthesis reported 95% CI in brackets
\* The authors also provide estimations for 18 pairs with certainty about onset dates.
<sup>U</sup> = unpublished, during peer review process
Source: own based of references

Considering the incubation period, if transmission occurs mainly inside the households the date of exposition is uncertain and restricts the estimations. Only 6 empirical studies provide figures about the time since exposition to illness onset (see Table 2).

**Table 2.** Incubation period of Covid-19. International evidence.

| Study | Region | N | Mean<br>-days- | SD<br>-days- |
| --- | --- | --- | --- | --- |
| Backer et al (2020) | travelers from and to Wuhan (China) | 88 | 6.4<br>[5.6-7.7] | 2.3<br>[1.7-3.7] |
| Li, Guan, Wu et al (2020) | China | 10 | 5.2<br>[4.1-7.0] | nr |
| Lauer et al (2020) | China excl Hubei | 181 | 5.5<sup>M</sup><br>[4.5-5.8] | nr |
| Linton et al (2020) | China excl Wuhan | 52 | 5.0<br>[4.2-6.0]* | 3<br>[2.1-4.5] |
| Zhang et al (2020) | China excl Hubei | 49 | 5.2<br>[1.8-12.4] | nr |
| Zhou et al (2020) | China excl Hubei | 65 | 5.33<br>5.0<sup>M</sup> | 3.3 |

M = median
\* the mean is around 5.6 with SD of 2.8 when inhabitants of Wuhan were included
Source: own based on references

## Materials and Methods

Between March 20, 2020 and May 8, 2020 36 cases were reported as positive for SARS-Cov-2 in Bahia Blanca city (Argentina). Local health authorities collected the date of illness onset and the date of exposure of each patient. As most of them emerged in clustered transmission it was possible to identify the infector-infectee pair in the chain.

From the total confirmed cases, 13 of them were imported and 23 were local cases. In addition, 4 of 23 local cases (17%) were asymptomatic up to May 8, 2020.

We have 17 observations of individual serial intervals emerged in 7 clusters of patients and 15 individual observations about the incubation period in patients who reported the date of exposition to their infector. Also, the date of exposition is probable in 3 cases, so we re-estimated the incubation period excluding those cases from the sample.

In most pairs (70%), the infectee is part of hospital staff and contracted the disease in workplace, in close contact with patients. For some of them (4 cases) the infector could not be clearly identified as more than one patient could have transmitted the virus. In those cases, we assigned the infector as the one exhibiting the closest period with the infectee’s illness onset. For that reason, following the procedure adopted by Nishiura et al (2020), we split the sample of pairs between certain and probable observations. The subsample of certain pairs has 13 observations.

We assumed the serial interval follows a Gamma distribution and time to symptoms is distributed as a Lognormal (Lesser et al, 2009). The parameters were estimated using the *fitdist* command from *fitdistrplus* package in R using the maximum likelihood estimator, accepted as the best method to estimate the time to event from patient data (Wallinga & Teunis, 2004; Bolker 2008). Confidence intervals for the median were obtained using the quantile matching estimator.

We also estimate the mean SI from the sample of patients assuming uncertainty in probable dates of symptoms onset. This requires a Bayesian approach, based on a numerical technique known as Markov Chain Monte Carlo (*mcmc*). This procedure estimates a posterior sample of SI distribution, which is a kind of average between prior beliefs of the distribution (e.g Gamma) and observed data. The Bayesian approach can be more accurate in diseases with less known dynamics.

## Results

In the sample of 17 infectee-infector pairs we found that 2 patients became infected during pre-symptomatic phase as their infector manifested symptoms after the exposition. Also, in the sample of cases with incubation dates, 1 case got infected from an asymtomatic infectious. Taking togheter, the proportion of transmission before symptoms onset or from asymtomatic cases is 16.6% (3 cases between 18).

As the number of observations is modest and parametric estimation methods have asymptotic properties, we also checked results with bootstrapping techniques (see Supplementary Materials).

Findings are presented in Table 3. The mean incubation period for symptomatic patients is 7.9 days (95% CI 4.6, 11.1) considering the sample of 15 cases patients and 7.5 days (95% CI 4.1, 10.9) if just the most certain cases (n=12) are considered. The median is 6.1 (95% CI: 4.1, 9.2) and 5.8 (95% CI 3.6, 9.3) respectively.

**Table 3.** Incubation and serial interval in a sample of symptomatic infections.

|  | Incubation period | Incubation period for most certain cases | Serial interval Parametric approach | Serial interval for most certain pairs Parametric approach | Serial interval full sample Bayesian approach | Serial interval for most certain pairs Bayesian approach |
| --- | --- | --- | --- | --- | --- | --- |
| Assumed distribution | Log normal | Log normal | Gamma | Gamma | Gamma** | Gamma** |
| Mean (1) | 7.86<br>[4.63-11.09] | 7.50<br>[4.11-10.89] | 6.82<br>[4.04-9.61] | 5.46<br>[2.84-8.08] | 6.88 | 6.12 |
| Standard deviation (2) | 6.38<br>[3.42-15.87] | 6.23<br>[3.21-16.99] | 5.86<br>[1.80-3.69] | 4.81<br>[2.48-13.12] | 5.89 | 4.55 |
| Median* | 6.1<br>[4.06-9.18*] | 5.76<br>[3.56-9.33*] | 5.24<br>[2.96-8.11*] | 4.13<br>[2.04-6.93*] | 6.4<br>[4.6-9.2] | 5.15<br>[2.95-7.50] |
| CV (2)/(1) | 0.81 | 0.83 | 0.86 | 0.88 | 0.86 | 0.74 |
| N | 15 | 12 | 17 | 13 | 17 | 13 |
| lnL | -43.31 | -34.19 | -49.21 | -34.83 | -52.52 | -37.96 |

\* CI for the median computed based on quantile matching estimator. Parameters differ only in the standard deviation estimation
\*\* it is the prior distribution, not the posterior
95% CI and 95% CrI in brackets, depending on parametric or Bayesian estimation
Source: own calculations

In addition, 97.5% of symptomatic cases will develop symptoms after 13.6 days from exposition (95% CI 10.7, 16.5). This results emerges from parametric boostraping. If re-sampling is non parametric, the figure decreases to 12.8 days (95% CI: 9.8, 15.9). In the reduced sample of most certain 12 cases is considered, 97.5% of cases will be ill after 14.5 days or 12.3 days, depending on boostraping method (parametric or non parametric. The upper limit of confidence interval increase to 17 days taking into account the results of the reduce sample. These estimations are useful to decide the extent of quarantine in exposed individuals. Even when 14 days seems an appropiate length, adding 3 extra days could be more effective to limit spread. The point estimation for the mean serial interval is 6.8 days (95% CI: 4.0-9.6). Considering only the most certain pairs, the mean serial interval is estimated at 5.5 days (95% CI: 2.8, 8.1). The estimated median serial intervals were 5.2 (95% CI: 3.0, 8.1) and 4.1 (95% CI: 2.0, 6.9) respectively. We found no substantive differences in the mean between the point estimations of parametric and Bayesian methods and also considering bootstraping (see Supplementary Materials). Nevertheless, Bayesian methods show higher mean and median for the SI in the reduced sample than the ones estimated without uncertainty. This finding narrows the gap between the SI and the incubation period.

Figure 1 plots the histogram of observations and fitted distributions. Estimations show better adjustment to observed data for the SI than for incubation period.

**Figure 1:**
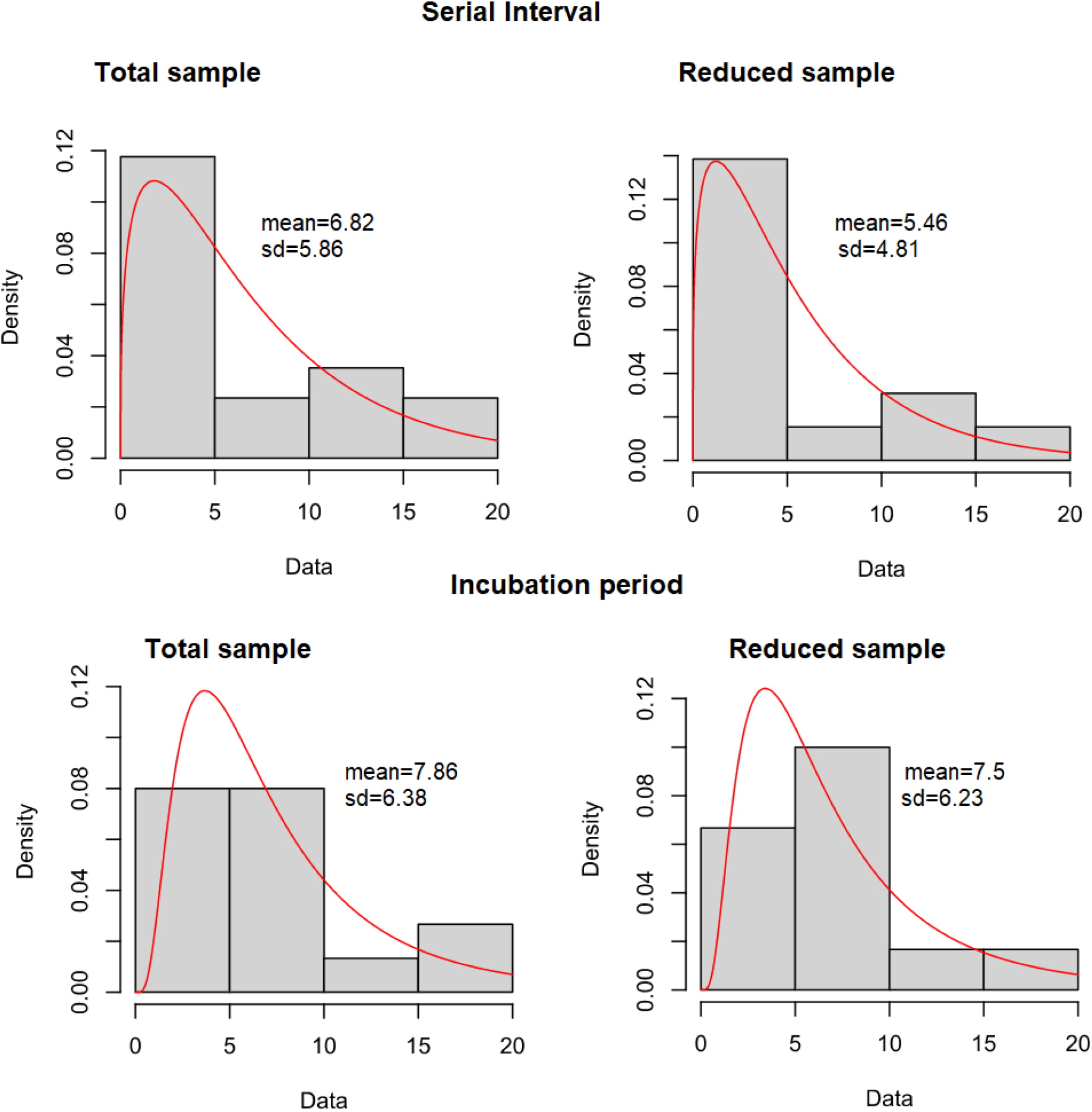
**Empirical and theoretical density of serial interval and incubation period distributions for total and reduced sample. Results from parametric estimations**

## Discussion

The mean incubation period of Covid-19 estimated for Bahia Blanca city seems longer than the one reported for Asian regions. That implies a slower propagation as long as there is no substantial pre-symtomatic or asymptomatic transmission. Anyway a note of caution must be posed, as the coefficient of variation is also higher in Bahia Blanca, warning that heterogeneity in local spreading is also higher than in Asian regions and/or the sample size is still low to get precise estimates.

The estimations for the serial interval of Covid-19 suggest that symptoms onset may take longer time to emerge between infectious, enlarging the effective reproductive number (conditional to the rate of growth of cases). Nonetheless, the SI seems quite close to the figures reported for Lombardy (Italy).

The gap between the incubation period and the SI seems similar than the one estimated in foreign regions; presymptomatic transmission can occur between 1 or 2 days before symptoms onset. That highlights the importance of contact tracing and timely isolation measures of patients’ close contacts.

The present study has some limitations. First, from the first 36 cases 33% of symptomatic cases are hospital staff for whom contagion is clearly during symptomatic phase and not before. If transmission becomes communitary, pre-symptomatic contagion could be higher than estimated one.

## Ethics Approval

Data collection and analysis of cases were conducted by the staff of the Epidemiology Department of the municipal government of Bahia Blanca. They are part of a continuing public health outbreak investigation. Thus data were considered exempt from institutional review board approval.

## Data Availability

Microdata about incubation dates and symptom onset can be delivered if requested

## Supplementary material

### Estimations with parametric and non parametric bootstrapping

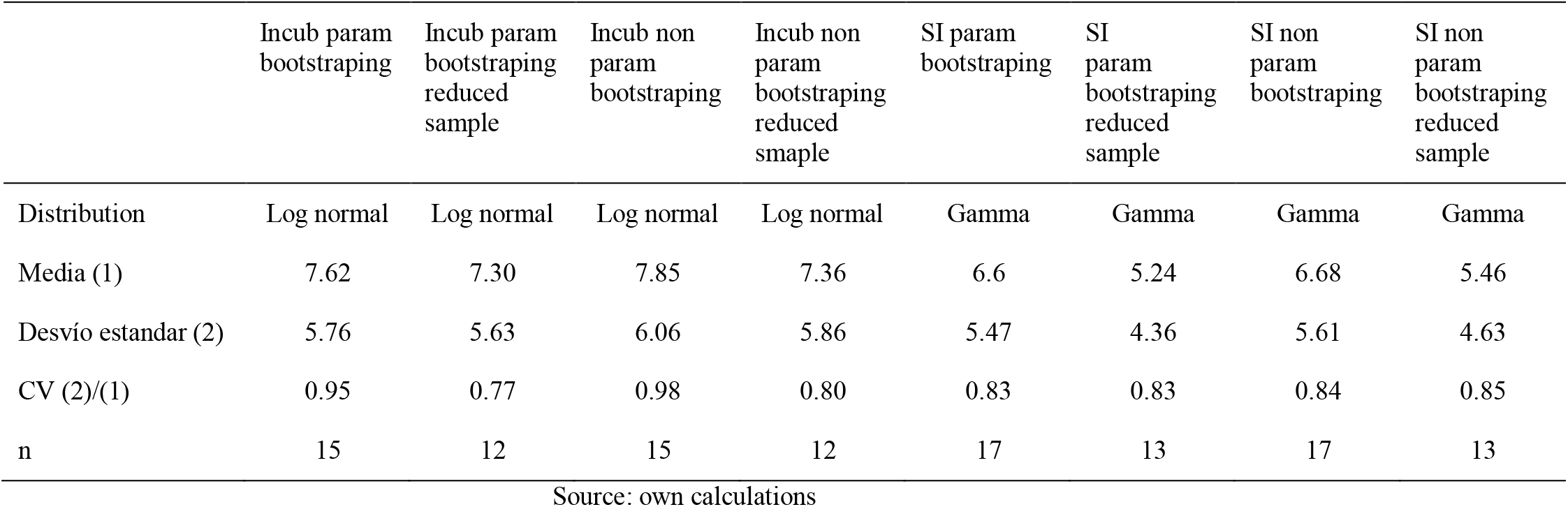

| Estimations with parametric and non parametric bootstrapping |  |  |  |  |  |  |  |  |
| --- | --- | --- | --- | --- | --- | --- | --- | --- |
|  | Incub param<br>bootstraping | Incub param<br>bootstraping<br>reduced<br>sample | Incub non<br>param<br>bootstraping | Incub non<br>param<br>bootstraping<br>reduced<br>smaple | SI param<br>bootstraping | SI<br>param<br>bootstraping<br>reduced<br>sample | SI non<br>param<br>bootstraping | SI non<br>param<br>bootstraping<br>reduced<br>sample |
| Distribution | Log normal | Log normal | Log normal | Log normal | Gamma | Gamma | Gamma | Gamma |
| Media (1) | 7.62 | 7.30 | 7.85 | 7.36 | 6.6 | 5.24 | 6.68 | 5.46 |
| Desvío estandar (2) | 5.76 | 5.63 | 6.06 | 5.86 | 5.47 | 4.36 | 5.61 | 4.63 |
| CV (2)/(1) | 0.95 | 0.77 | 0.98 | 0.80 | 0.83 | 0.83 | 0.84 | 0.85 |
| n | 15 | 12 | 15 | 12 | 17 | 13 | 17 | 13 |

Source: own calculations

### Q-Q Plots of serial interval and incubation period fits

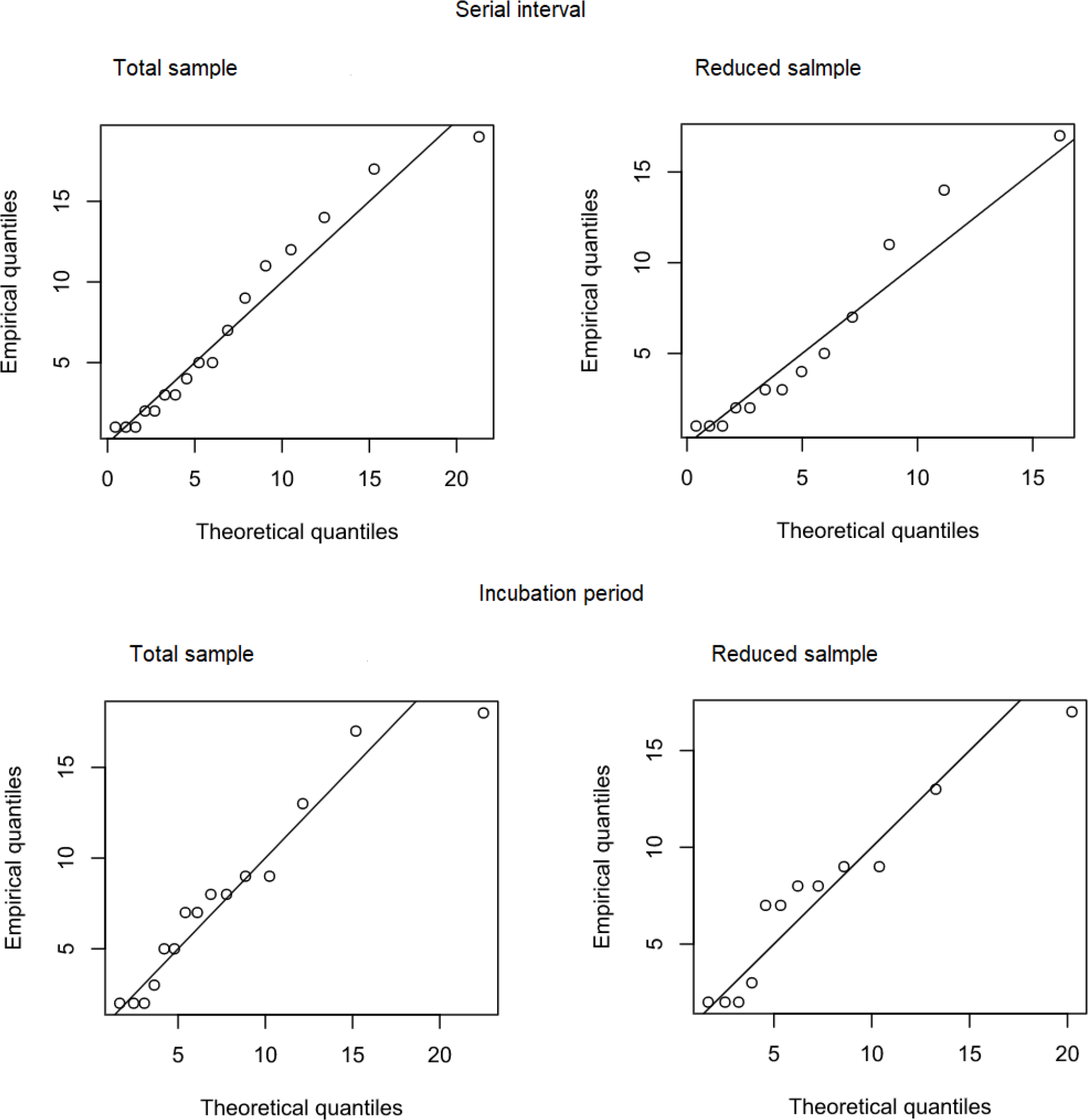

